# Mutations in emerging variant of concern lineages disrupt genomic sequencing of SARS-CoV-2 clinical specimens

**DOI:** 10.1101/2021.06.01.21258181

**Authors:** Kevin S. Kuchinski, Jason Nguyen, Tracy D. Lee, Rebecca Hickman, Agatha N. Jassem, Linda M. N. Hoang, Natalie A. Prystajecky, John R. Tyson

## Abstract

Mutations in emerging SARS-CoV-2 lineages can interfere with the laboratory methods used to generate high-quality genome sequences for COVID-19 surveillance. Here, we identify 46 mutations in current variant of concern lineages affecting the widely used laboratory protocols for SARS-CoV-2 genomic sequencing by Freed et al. and the ARTIC network. We provide laboratory data showing how three of these mutations disrupted sequencing of P.1 lineage specimens during a recent outbreak in British Columbia, Canada, and we also demonstrate how we modified the Freed et al. protocol to restore performance.

## Background

Genomic sequencing of SARS coronavirus-2 (SARS-CoV-2) has played a crucial role in managing the on-going COVID-19 pandemic. This is especially true for variant of concern lineages that have emerged globally since December 2020 [1-6]. Genomic sequencing has been instrumental in detecting and characterizing these lineages as they emerge, tracking their global spread, and identifying local cases to control transmission.

Due to low quantities of viral genomic material in typical clinical specimens, the SARS-CoV-2 genome must be amplified for high-throughput sequencing. This is commonly done by amplicon sequencing. This approach involves multiplexed PCR with numerous primers pairs tiled across the viral genome [7-9]. The performance of these primer schemes can be impacted by mutations associated with emerging viral lineages. These mutations create thermodynamic instabilities between primer oligonucleotides and genomic template material, interfering with amplification. This causes drop-outs in sequencing coverage and hinders the collection of complete, high-quality viral genomes.

### Amplicon drop-outs in P.1 lineage specimens using the Freed *et al*. primer scheme

The British Columbia Centre for Disease Control’s Public Health Laboratory (BCCDC PHL) currently uses the 1200 bp amplicon scheme by Freed *et al*. for genomic sequencing of SARS-CoV-2 clinical specimens [8]. While investigating a local P.1 lineage outbreak in March and April 2021, the BCCDC PHL observed significantly reduced depth of coverage across three amplicons covering parts of the orf1ab and spike genes (Figure 1). This suggested that mutations in the P.1 lineage had occurred in primer sites targeted by the Freed *et al*. amplicon scheme.

**Figure 1:**
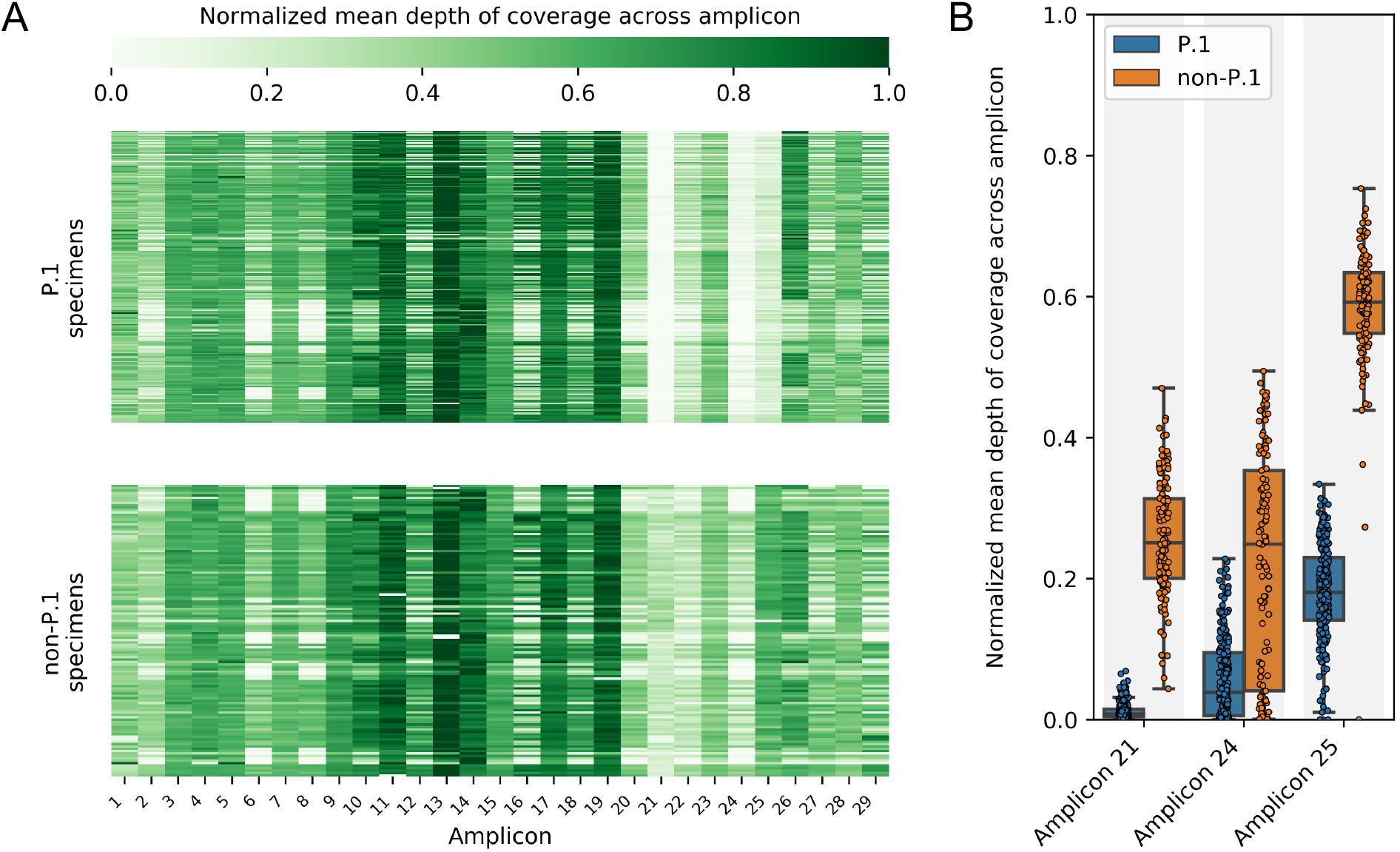
Predominant mutations in the P.1 lineage reduce sequencing depth across three amplicons in the Freed *et al*. primer scheme. Mean depths of coverage across all amplicons were calculated for 344 SARS-CoV-2 clinical specimens collected during March and February 2021 in British Columbia, Canada. Mean depths of coverage were normalized to the mean depth of coverage of each library’s most deeply sequenced amplicon. A) Mean depths of coverage for all amplicons in all specimens are visualized in a heatmap stratified by P.1 and non-P.1 lineage. Amplicon 21, 24, and 25 drop-out is apparent in P.1 lineage specimens. B) Distributions of normalized mean depths of coverage are shown for the three amplicons impacted by widespread mutations in the P.1 lineage. Differences between P.1 and non-P.1 lineage specimens were significant for all three amplicons (p-values <0.00001, Wilcoxon rank sums test).

We used a previously published bioinformatic pipeline called PCR_strainer to search P.1 lineage genome sequences for mutations in sites targeted by the Freed *et al*. primers [10]. We analyzed 907 local P.1 lineage sequences generated by the BCCDC PHL before April 7, 2021. The consensus and coverage metrics were generated from short-read reference guided sequencing assembly and analysis using an OICR fork of the COG-UK nextflow pipeline and the ncov-tools package [11,12] together with lineage assigned using pangolin [13]. We also analyzed 1,634 global P.1 lineage sequences collected and submitted to GISAID before April 7, 2021. We identified three primer site variants that were widespread locally and globally, appearing in over 98% of P.1 sequences analyzed (Table 1). These primer site variants affected the same amplicons identified above with reduced depths of coverage in P.1 lineage clinical specimens.

**Table 1:**
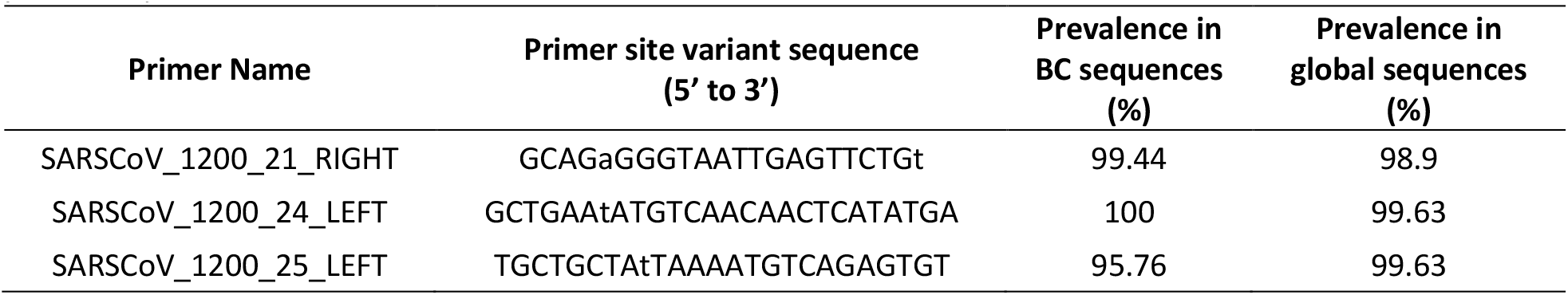
Nucleotide mismatches between Freed *et al*. amplicon sequencing primers and SARS-CoV-2 P.1 lineage. Global P.1 lineage sequences were downloaded from GISAID, comprising 1,634 complete, high-coverage sequences collected and submitted before April 7, 2021. Sequence details and submitting laboratories, who we gratefully acknowledge for their contributions, are provided in Supplemental 1. British Columbia (BC) P.1 lineage sequences were generated before April 7, 2021 by the British Columbia Centre for Disease Control’s Public Health Laboratory and included 907 sequences. Primer site variants were identified using the PCR_strainer pipeline with default parameters. Primer site variants were only reported if they appeared in at least 5% of local or global sequences (excluding low coverage BC sequences). Primer site variant sequences are provided in ‘oligo sense’, *i*.*e*. the reverse complement of the primer site and the sequence that the perfectly identical primer would have for the targeted location. Lower case bases in the primer site variant sequences indicate mismatches with the default primer sequence.

One of these primer site variants contained 2 mismatches in the reverse primer for amplicon 21. One mismatch was located at the crucial 3’ end of the primer, which was reflected in amplicon 21 having the greatest reductions in mean depth of coverage (Figure 1B). The other two primer site variants we identified contained single mismatches with the forward primers for amplicons 24 and 25. These mismatches were located closer to the 5’ ends of these primers, so their observed impact on sequencing depth was less easily predicted. This demonstrates that the impact of mismatches identified *in silico* cannot be inferred from position alone and must be verified with laboratory data.

### Improving sequencing depth for problematic amplicons

To address drop-out across these problematic amplicons, we designed supplemental primers for the Freed scheme based on the primer site variant sequences identified by PCR_strainer. We called these primers 21_right_P.1, 24_left_P.1, and 25_left_P.1, and they had the following sequences: 5’-GCAGAGGGTAATTGAGTTCTGT-3’, 5’-GCTGAATATGTCAACAACTCATATGA-3’, and 5’-TGCTGCTATTAAAATGTCAGAGTGT-3’ respectively. These primers were spiked into default Freed *et al*. primer pools at the same molarity as the other primers. We sequenced 24 P.1 lineage clinical specimens with spiked and default primer pools, then we used these paired data to calculate improvements to sequencing depth across problematic amplicons (Figure 2A). The spike-in primers significantly improved mean depths of coverage for amplicons 21 and 25 (Figure 2A). Depth of coverage across amplicon 24 was not improved, however, so we also tried 24_left_P.1 spiked-in at two- and four-times molarity (Figure 2B). Increasing the spike-in primer concentration significantly improved mean depth of coverage for amplicon 24, with four-times molarity providing the greatest improvement.

**Figure 2:**
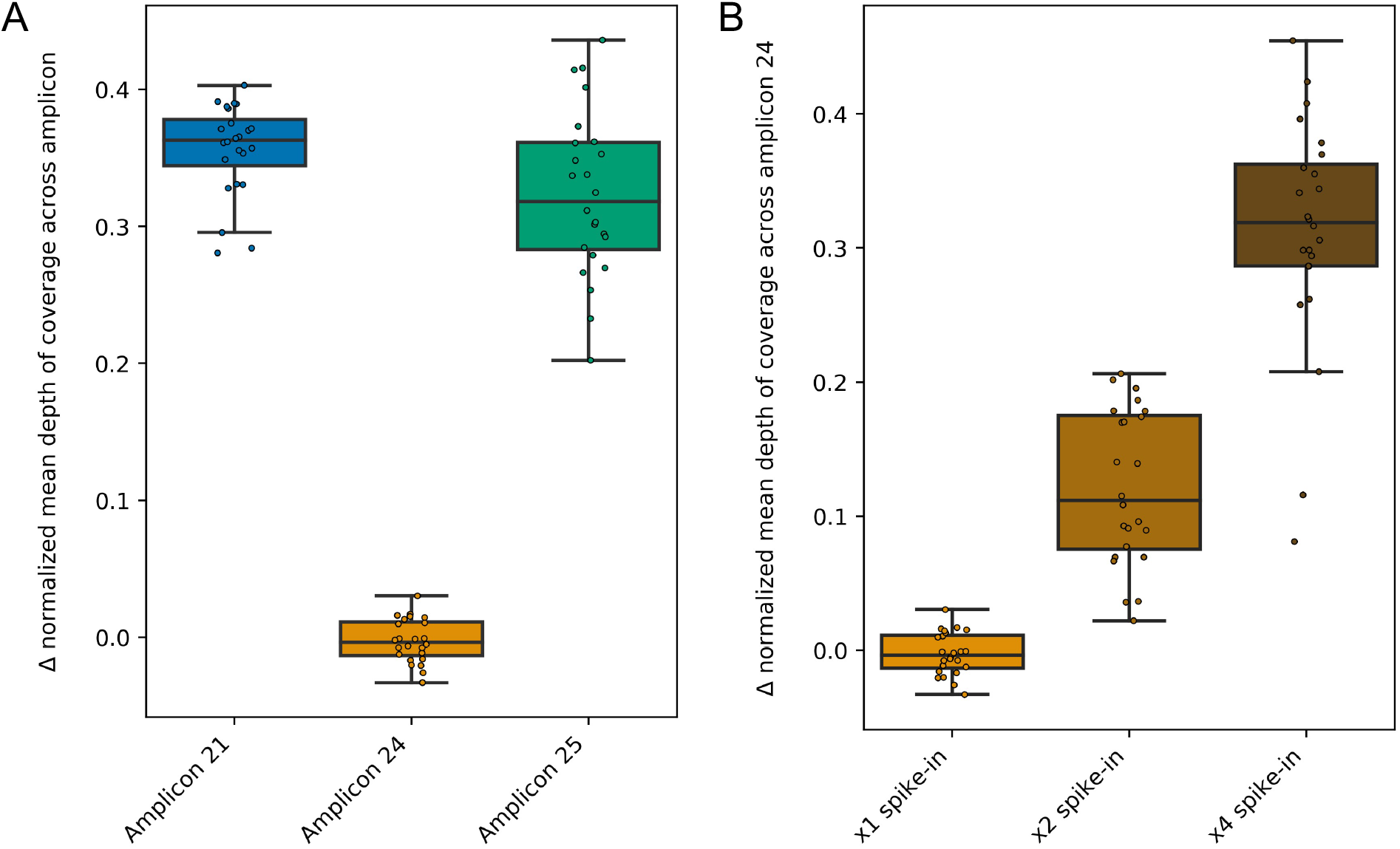
Amplicon drop-out is corrected by spiking updated primers into Freed *et al*. primer scheme. Updated primers were designed for problematic amplicons based on the primer site variant sequences identified by PCR_strainer in Table 1. Freed primer pools with and without these updated primers were used to sequence 24 P.1 lineage clinical specimens. Mean depths of coverage across each amplicon were calculated and normalized to each library’s most deeply sequenced amplicon. These paired data were used to calculate changes in normalized mean depths of coverage across amplicons. A) Depth of coverage was significantly improved for amplicons 21 and 25 by spiking the updated primers into the Freed pool at one-times molarity (p-values < 0.00001, paired sample T-test). B) Depth of coverage across amplicon 24 was significantly improved when the updated primer was spiked into the Freed pool at two- and four-times molarity (p-values < 0.00001, paired sample T-test).

Based on these results, we prepared Freed scheme primer pools with the supplemental primers spiked-in. Primers 21_right_P.1 and 25_left_P.1 were spiked-in at one-times molarity, and primer 24_left_P.1 was spiked-in at four-times molarity. Using these spiked primer pools, we sequenced 347 clinical specimens (Figure 3A). For amplicons 21 and 24, mean depths of coverage for P.1 lineages exceeded non-P.1 lineages or were on par with them. Depths of coverage across amplicon 25 for P.1 lineages still trailed non-P.1 lineages but were a significant improvement.

**Figure 3:**
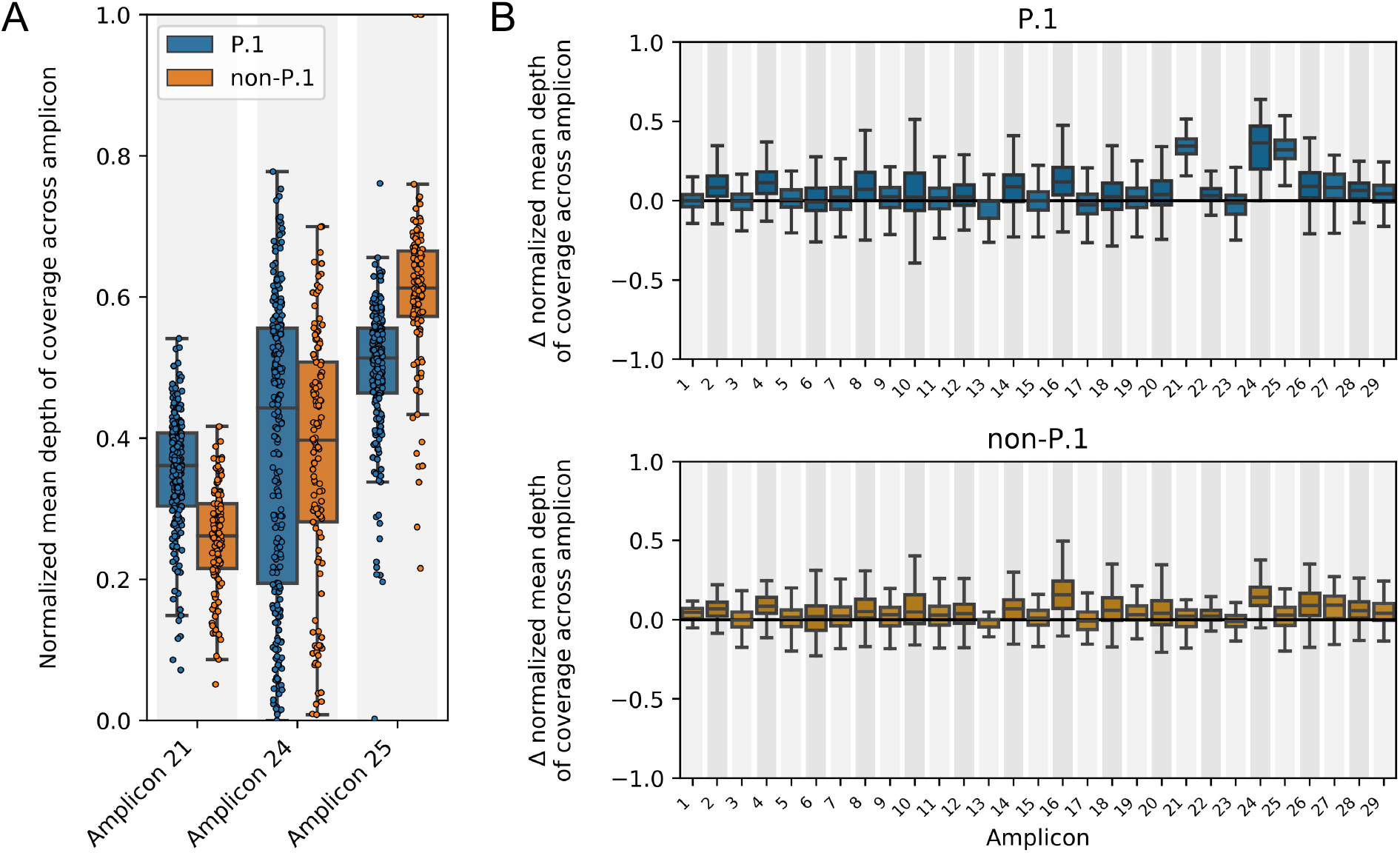
Spike-in primers improved depth of sequencing for amplicons impacted by dominant mutations in P.1 lineages without affecting other amplicons or other lineages. Freed primer pools were prepared with the updated primers spiked-in at one-times molarity (amplicon 21 and 25) or four-times molarity (amplicon 24). Mean depths of coverage across each amplicon were calculated and normalized to each library’s most deeply sequenced amplicon. A) 347 clinical specimens were sequenced with the updated primer pool. Mean depths of coverage were compared between P.1 and non-P.1 lineage specimens across amplicons impacted by widespread mutations in the P.1 lineage. B) 340 specimens sequenced using the updated primer pool were compared to previous results generated without the spike-in primers. No detrimental effects from the spike-in primers were observed for other amplicons or lineages.

Previous sequencing results using the default Freed pool were available for 340 of these clinical specimens. We used these paired data to calculate differences in sequencing depth between the default primer pool and our spiked primer pool (Figure 3B). We did not observe any detrimental impact from our spike-in primers on depth of coverage for other amplicons or non-P.1 lineages.

### Mutations in other variant of concern lineages impacting Freed *et al*. and ARTIC version 3 primer sites

We expanded our *in silico* analysis with PCR_strainer to assess the Freed *et al*. primer scheme against recent global sequences from all current variant of concern lineages (B.1.1.7, B.1.351, P.1, and B.1.617+) [1-6]. We also performed this analysis on the widely used and commercially supplied ARTIC version 3 primer scheme [9].

We identified 46 primer site variants affecting both protocols (Table 2). The B.1.1.7 lineage had the fewest primer site variants (n=5), followed by the B.1.351 lineage (n=9), followed by the B.1.617+ and P.1 lineages (both n=16). Many of these primer site variants were predominant globally within their lineage, as would be expected, with 12 of them being present in at least 90% of their lineage’s sequences. The ARTIC version 3 primer scheme was impacted by 34 primer site variants while the Freed *et al*. scheme was impacted by 12. We attribute this difference to the smaller amplicon sizes of the ARTIC version 3 protocol. This highlights an important implication of amplicon size when designing primer schemes for amplifying viral genomes. Longer amplicons require fewer primers, creating fewer opportunities for mutations to disrupt an assay. On the other hand, more genome coverage is lost with longer amplicons when a mutation impacts a primer site. This can affect a primer scheme’s longevity, and it should be balanced alongside the benefits of shorter amplicons for amplifying fragmented template material.

**Table 2:**
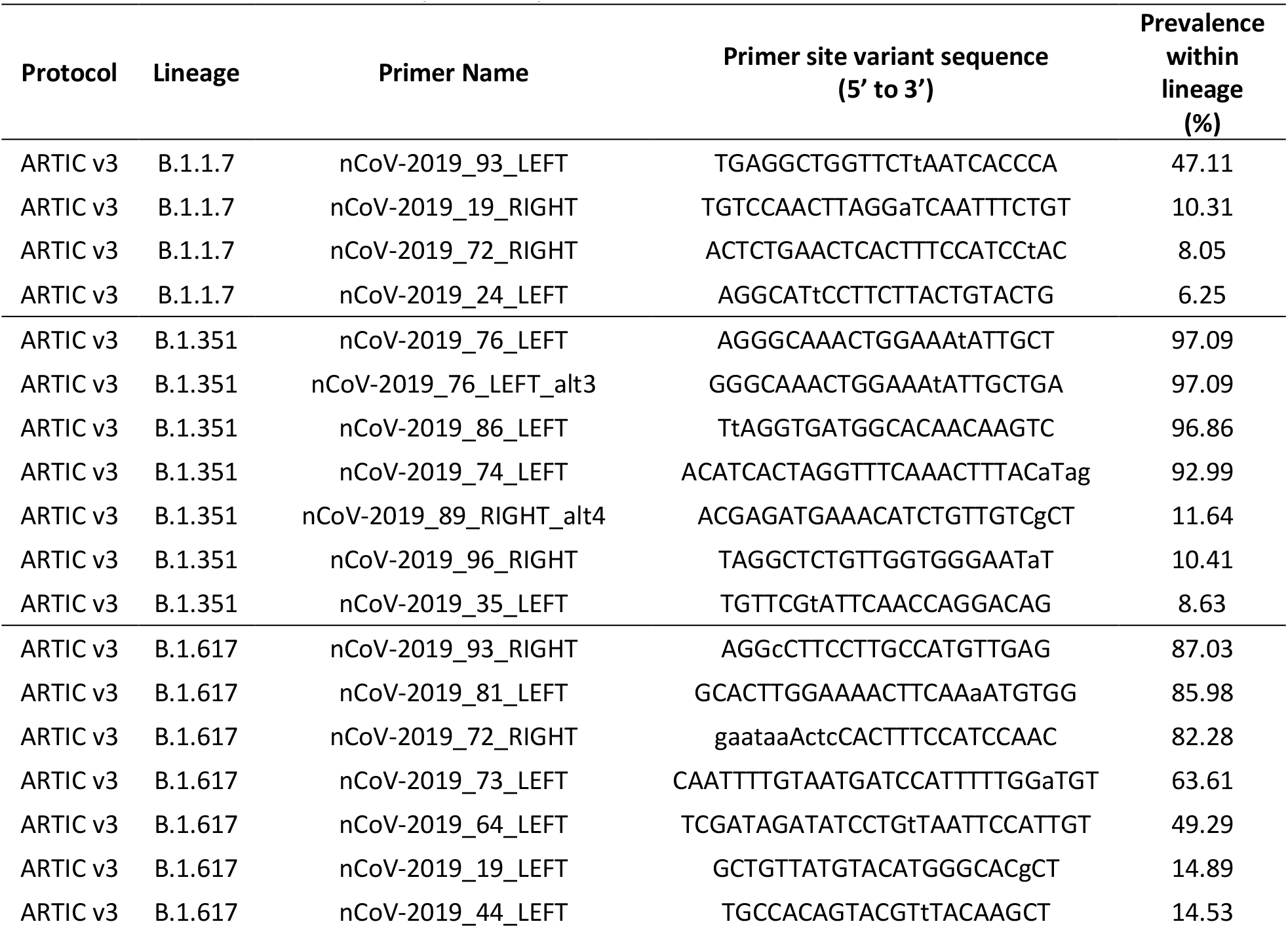
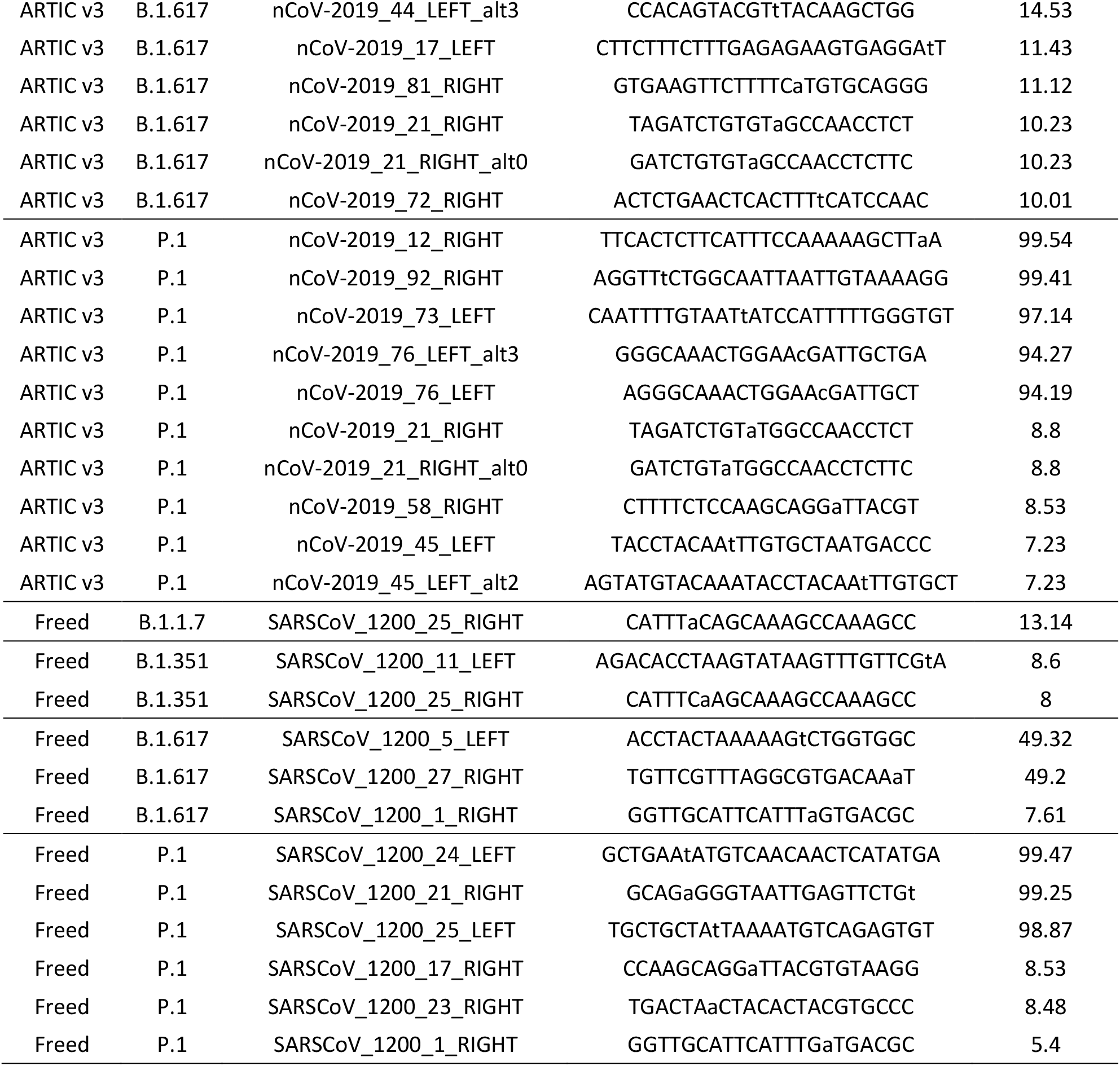
Nucleotide mismatches between SARS-CoV-2 variant of concern lineages and sequencing primers from ARTIC version 3 and Freed *et al*. protocols. Global sequences from variant of concern lineages were downloaded from GISAID, comprising 8,353 B.1.1.7 sequences, 3,025 B.1.351 sequences, 6,664 B.1.617+ sequences, and 9,202 P.1 sequences. All sequences were complete, high-coverage, and submitted before May 22, 2021. For lineages B.1.351, B.1.617+, and P.1, sequences were from specimens collected after April 1, 2021. Due to excess availability, sequences from lineage B.1.1.7 were from specimens collected after May 7, 2021. Sequence details and submitting laboratories, who we gratefully acknowledge for their contributions, are provided in Supplemental 2. Primer site variants were identified using the PCR_strainer pipeline with default parameters. Primer site variants were only reported if they were present in at least 5% of sequences from their lineage. Primer site variant sequences are provided in ‘oligo sense’, i.e. the reverse complement of the primer site and the sequence that the perfectly identical primer would have for the targeted location. Lower case bases in the primer site variant sequences indicate mismatches with the default primer sequence.

## Conclusions

This study demonstrates the importance of monitoring emerging viral lineages for mutations that might disrupt clinical sequencing and confirming their impact with laboratory data. Using PCR_strainer, we quickly identified three widespread mutations in the P.1 lineage of SARS-CoV-2 and designed successful spike-in primers for the Freed *et al*. primer scheme. We also used PCR_strainer to identify numerous widespread primer site mutations in variant of concern lineages that could impact the popular Freed *et al*. and ARTIC version 3 protocols. Our results suggest that extensive updates for widely used amplicon sequencing schemes are necessary immediately, and that primer schemes will have to evolve alongside SARS-CoV-2. Our results will be useful in the short-term for designing these updates. In the long-term, our combination of PCR_strainer analysis and laboratory validation provides a useful approach for maintaining SARS-CoV-2 clinical sequencing protocols.

## Data Availability

Laboratory protocols cited where relevant. Details and accession numbers of GISAID sequences analyzed are included as supplementals.

## Acknowledgements

We thank the dedicated staff at the BCCDC PHL for processing and sequencing SARS-CoV-2 clinical specimens, especially the Molecular Microbiology and Genomics program for optimizing genomics methods and the Bacteriology and Mycology program for routine sequencing of clinical specimens. We also thank the analytical staff for routine bioinformatic analysis, and Jared Simpson and members of the Simpson lab for provision of analysis and reportering software. This work was supported in part by the Canadian COVID genomics network (CanCOGeN).

## Author Contributions

KK wrote the manuscript under the direction of NP and JT. The study was conceived by KK, NP, AJ, TL, and JT. Laboratory work was performed by JN with support from TL and RH and under the direction of NP, LH, and JT. Bioinformatic analysis and data analysis was performed by KK and JT. All authors reviewed the manuscript and provided edits.

